# Investigating the impact of lived experience stories on self-harm, mood, and help-seeking intentions: an experimental study

**DOI:** 10.1101/2024.04.25.24306405

**Authors:** Jennifer Ferrar, Lizzy Winstone, Ian Penton-Voak, Lucy Biddle, Paul Moran, Lydia Grace, Becky Mars

## Abstract

**Objective:** To investigate the psychological impact of variations in help-seeking messages contained in lived experience stories about self-harm.

**Method:** In an online experiment, individuals with a recent history of self-harm, were randomised to read stories that either mentioned: i) self-help strategies, ii) seeking help from informal and formal sources, or iii) did not mention help-seeking. Help-seeking intentions, mood, entrapment, and expectations of future self-harm was measured, and participants provided feedback on the stories.

**Results:** There was limited evidence for an effect of story type on future help-seeking intentions and stronger evidence for an effect of story type on negative affect. Participants in the “Self-help” condition reported lower negative affect after reading the stories compared to participants in the “No help” condition (Mean difference = -3.97, 95% CI -7.72 to -0.22, p = .04) and the “Informal/formal” help condition (Mean difference = -3.70, 95% CI -7.55 to 0.14, p =.06). A key criticism of the stories was that they were unrelatable, but this sentiment was less prevalent among those in the “No help” condition. Key positives were that the stories included a realistic but hopeful outlook of recovery (less prevalent in the “Informal/formal help” condition) and were supportive (less prevalent in the “No help” condition).

**Conclusion:** While the inclusion of self-help strategies in a lived experience story reduced its impact on negative affect, the inclusion of self-help strategies or seeking help from others did not encourage future help-seeking intentions. Making stories relatable, authentic, and providing multiple strategies for support might be key to encourage help-seeking, but more research is needed.

## Introduction

Individuals experiencing self-harm or suicidal thoughts may seek support via the internet. Self-harm related internet use can be both helpful and harmful (1,2). Self-harm related internet content can foster a sense of community, thereby reducing feelings of loneliness and isolation, (3–6) and provide practical advice and moral support on how to manage self-harm (4,7–10). In addition, self-harm related internet use provides an alternative for individuals who are not yet ready, or may be unwilling, or unable to access support offline (11–14).

However, there is also a link between self-harm related internet use and higher levels of suicidal intent (12,15,16), although the direction of effects is not clear. Due to a lack of regulation, content encouraging self-harm is easily accessible online (2,17,18). In addition, content or environments intended to discourage self-harm could unintentionally reinforce self-harm behaviour, for example via normalisation or competition (18,19). Further, the categorisation of self-harm content as either harmful or helpful is not straightforward. A recent systematic review illustrates the complex nature of engagement with self-harm content. Whether the content negatively or positively affects the individual engaging with it depends on the content, the individual, and the time and space in which it is encountered (2).

The internet provides a wide range of self-harm content including information about self-harm behaviour, self-help strategies, lived-experience stories, images of self-harm, signposting to help, and dialogue (2,13,18,20,21). They all come with limitations, for example, research suggests that information-giving can often be impersonal, disengaging, and “insufficient in moments of crisis” and that signposting can often be redundant as many searching for help online are doing so because of barriers to engaging with signposted services (13). The impact of self-harm images such as scars can be mixed, reminding some of recovery while triggering others (2). Among those who access self-harm related internet content, there seems to be a preference for real-time help (e.g., “live chat”), self-help tools, and interaction with others, particularly via lived-experience content (13,22).

Lived-experience stories are often used on formal help sites as they are viewed as personal and engaging and can validate crisis for readers, while also offering reassurance that they are not alone in their struggle and that recovery is possible (13,22,23). However, there are several components which might influence whether a lived-experience story is perceived as helpful, unhelpful, or potentially harmful. Stories appearing on forums, even those that are run by trained moderators, might be triggering for some people, for example by including details of methods, injury, or graphic imagery. Stories appearing on websites run by charities and healthcare organisations usually include a personal narrative of help-seeking and recovery. However, the evidence supporting that these encourage help-seeking in the reader is limited (22,23). Further, the careful regulation of content might be perceived as overly censored and lead some to search for unrefined content in covert and riskier spaces (2). Even unregulated content posted by a lay individual is curated in a different respect, for example on social media platforms where users are often presenting themselves in an idealized way (24,25).

The present study is part of a mixed-methods project which aimed to investigate how variations in help-seeking messages contained within lived experience stories are interpreted by and psychologically impact those with self-harm experience. Findings from the qualitative component (focus groups) demonstrated that although it is a complex relationship, individuals with recent self-harm experience could positively engage with lived experiences stories, for example if they reflected a wide range of personal journeys and avoided the use of stigmatizing language (23). The present study aimed to gather additional evidence on the effect of online lived-experience stories using an experimental design.

More specifically, we sought to investigate the psychological impact of stories with different types of help-seeking messages, including those mentioning self-help strategies, informal/formal help-seeking (i.e., friends, family, or professionals), and no help-seeking. We sought to assess if the type of help-seeking referenced in a lived experience story had an impact on help-seeking intentions, mood, expectations of future self-harm behaviours, as well as psychological factors implicated within the suicidal pathway, such as sense of entrapment (i.e., lack of escape from unbearable thoughts, feelings, situations) (26,27).

## Methods

This paper focuses on the quantitative component of a mixed methods study (for the qualitative component, see Winstone et al., 2023) which explored how people with self-harm experience and interpret lived experience accounts as a means of support. The protocol (https://osf.io/qtgvp) was pre-registered ahead of data collection (https://osf.io/cesda).

### Participants

The study was advertised by the research team and mental health charities on their webpages, social media accounts and in newsletters sent out to individuals who signed up to hear about regular research/charity activity. The study was also advertised on self-harm/mental health specific forums, on Reddit, RecoverYourLife, and the National Self Harm Network website. The study was hosted online: after providing consent and completing a self-administered baseline questionnaire, participants were randomly assigned to one of three story conditions 1) self-help strategies, 2) informal/formal help-seeking, 3) no help-seeking.

We conducted an a priori power analysis (G*Power v3.1.9.7) (28) which determined that a sample of 246 participants would provide adequate (80%) power for our main analysis (one-way ANOVA) to detect a small-medium effect size (f = 0.2) for our primary outcome (differences in help-seeking intentions) among the three story groups.

Ethical approval was granted by the University of Bristol School of Psychological Science Research Ethics Committee (REF: 10504). The study was conducted according to the revised Declaration of Helinski (2013) and the 1996 (ICH Guidelines for Good Clinical Practice E6(R2). To be eligible for the study, participants had to be aged 16 years or older, fluent in English, currently resident in the UK, and have engaged in self-harm within the past year. Before beginning the study, participants had to provide informed written consent. After taking part, participants were offered the opportunity to enter in a prize draw to win one of four £40 shopping vouchers.

### Setting

The task was programmed using Gorilla Experiment Builder (https://gorilla.sc/) (29). Participants could access the ask from any device that allowed them to access any web browser.

### Materials

Three template stories were used in this study (see Appendix S1). The same three templates were presented to all participants but a portion of text was modified (removed and/or inserted) to reflect the different experimental conditions: Self-help strategies, informal/formal help-seeking, and no help-seeking. The template stories were chosen from an initial shortlist of ten real-life stories curated by BM. These were selected from a variety of online sources including the websites of third-sector organisations and healthcare organisations, web-based forums, and personal blogs. This approach was used to ensure that they were not solely representative of the more polished stories that often appear on formal help sites. For ethical reasons, stories were excluded if they encouraged self-harm, described self-harm methods in detail or did not have a recovery-oriented/hopeful outlook. Each of the ten stories was independently ranked (in order of preference) and discussed by BM and three of the study authors (JF; IPV; LB). Three lived experience stories were then chosen following the ranking and discussion to be used as the template stories. The only key difference across the experimental conditions was whether text concerning self-help strategies, informal/formal help-seeking, or no help-seeking (but positive/hopeful message) was inserted into the stories. These inserts were drawn from the original ten stories selected by BM.

### Measures

#### Participant characteristics (assessed at baseline)

Participants reported their age in years, their gender, sexual orientation, and ethnicity. To measure help-seeking history, participants completed a modified version of the Actual Help-Seeking Questionnaire (AHSQ)(30,31). They also answered questions on self-harm thoughts (past year frequency), passive and active suicidal thoughts (in lifetime and recency), non-suicidal self-harm (in lifetime, recency, and past year frequency) and suicide attempts (in lifetime and recency). For further details, please refer to the protocol (https://osf.io/qtgvp).

#### Outcome measures (assessed post randomisation)

##### Primary outcome: Help-seeking

To measure help-seeking intentions, participants completed a modified version of the General Help Seeking Questionnaire (GHSQ) (32).

##### Secondary outcomes: Affect, Entrapment, and Future self-harm

Participants completed The Positive and Negative Affect Schedule (PANAS) (33). They completed the 4-item Entrapment Scale Short Form (E-SF) (34). They also answered two questions about future self-harm, rated on a 5-point scale. Firstly, how likely they would be to hurt themselves on purpose in the future after reading the stories, and secondly, how likely it was that they could see a time in the future without self-harm.

##### Opinions of the lived experience stories

Participants were asked to provide open-ended feedback on the lived experience stories presented to them during the study. Specifically, they were asked, “Was there anything you found helpful/unhelpful about the stories? If so, please explain.”

##### Attention check

To ensure that only data from participants who were paying attention during the study was included in the analysis, one of the items among the outcome measures asked participants to select 1 on a scale from 0 to 4. Participants who selected a number other than 1, had their data removed prior to the analysis (35).

### Procedure

Participants completed the questionnaires on demographics, current/past self-harm thoughts and behaviours, and help-seeking history. They were then randomised, using Gorilla’s randomisation feature, to one of the three conditions (i.e., self-help, informal/formal help-seeking, or no help-seeking). Participants were presented with and asked to read the three lived experience stories, with amended text based on the condition that they were assigned to. They then completed the outcome measures in the following order: i) free text opinions of the lived experience stories, ii) mood, iii) entrapment, iv) thoughts of self-harm/suicide, and v) help-seeking intentions.

### Analysis Plan

#### Data screening

To maintain data quality, each participant’s data was screened against the following criteria prior to analysis: each participant must have completed the entire study, and correctly answered the eligibility questions and the attention check question within the experiment. The data was assessed for normality using skewness and kurtosis statistics, and that it met the assumptions for ANOVA testing.

#### Data analysis

Descriptive statistics were used to summarise the participant characteristics and outcome measures in each group. An ANOVA was used to test if the type of help-seeking mentioned in the lived experience story influenced the primary outcome of interest (i.e., total score on the GHSQ). A series of ANOVAs was used to test if the type of help-seeking mentioned in the lived experience story influenced the secondary outcome measures (i.e., total scores on PANAS, E-SF and the two items assessing future self-harm). Where the type of help-seeking mentioned in the stories impacted any of the outcomes, post-hoc pairwise comparisons were conducted (with corrections applied for multiple comparisons).

This study was powered to detect a main effect of type of lived experience story, therefore any interactions were exploratory. Where any of the analyses indicated a main effect of type of lived experience story on the outcome measures, then story type x self-harm history ANCOVAs were used to assess if the influence of type of lived experience story on the outcome measures differed by self-harm history. The self-harm history variable was defined as the recency of self-harm behaviour in the past year. In cases where a participant had a history of both NSSH and attempting suicide, the recency score (indicating “either in the past week” or “more than a week ago, but in the past year”) for whichever behaviour was more recent was used. Where any interactions existed, they were explored in post-hoc analyses using t-tests. All analyses were run both unadjusted and adjusted for age, gender, and a continuous variable of help-seeking history (i.e., total score on the AHSQ).

Findings from the qualitative component (focus groups) of this mixed-method project suggested that lived experience stories can have a positive impact on readers, but that this effect may be moderated by age (23). In the protocol, we had specified that exploratory analyses would be guided by the qualitative results. Therefore, we have included a tertiary analysis, exploring interactions between story type and age on any primary and secondary outcomes where main effects were found.

Participant opinions on the lived experience stories were analysed using content analysis. After reviewing the data, specifically the length and richness of responses, it was decided that instead of conducting thematic analysis (as originally specified in the protocol), it would be more appropriate to conduct content analysis. Responses were first open coded by JF and JS (see acknowledgments). JF then refined the codes before LW coded 10% of the responses, adding additional codes when they felt it was appropriate. Intercoder reliability (ICR) was then assessed and calculated to be around 99%. JF then refined the codes once more.

## Results

### Participants

#### Demographics

Data collection ceased when there were complete datasets from 246 participants. One participant was identified as completing the study twice and removed from the dataset. Of the remaining 244 participants, data cleaning procedures identified two participants who failed the attention check and six who did not meet the eligibility criterion of having hurt themselves on purpose in the past year. Therefore, data from 238 participants was included in the analysis (97% of our target sample size). Participants were primarily female (79%), white (87%) and were aged between 16 and 59 years old (M = 23.2, SD = 9.4). Table 1 provides further details on participant demographics across the experimental conditions, as well as a breakdown by experimental condition. The experiment ran for 28 weeks (from 26^th^ March to 8^th^ October 2022).

**Table 1.** Participant demographics.

|  | *All*<br>*(n=242)* |  | *Self-help*<br>*(n=83)* | *Informal/fo*<br>*rmal help*<br>*(n=75)* | *No help*<br>*(n=80)* |
| --- | --- | --- | --- | --- | --- |
| **Age $M(SD)$** | 23.2(9.4) |  | 23.4(9.6) | 21.6(8.2) | 23.8(10.2) |
| **Gender** | **n** | **%** | **%** | **%** | **%** |
| Female | 188 | 79 | 79 | 73 | 84 |
| Male | 25 | 11 | 9 | 16 | 7 |
| Prefer to self-describe<sup>1</sup> | 20 | 8 | 10 | 9 | 6 |
| Prefer not to say | 5 | 2 | 3 | 1 | 2 |
| **Ethnicity** | **n** | **%** | **%** | **%** | **%** |
| Arab | 2 | 1 | 1 | 1 |  |
| Asian/Asian British | 4 | 2 | 1 | 4 |  |
| Black/African/Caribbean/Black British | 6 | 2 | 4 |  | 4 |
| White | 207 | 87 | 81 | 87 | 93 |
| Mixed/multiple ethnic groups | 17 | 7 | 12 | 7 | 2 |
| Other ethnic group<sup>2</sup> | 1 | <1 |  | 1 |  |
| Prefer not to say | 1 | <1 |  |  | 1 |
| **Sexual orientation** | **n** | **%** | **%** | **%** | **%** |
| Asexual or Aromantic (attracted to no genders) | 11 | 5 | 5 | 5 | 4 |
| Bisexual or pansexual (attracted to all genders) | 88 | 37 | 36 | 39 | 36 |
| Gay or lesbian (attracted to people of the same gender) | 23 | 10 | 12 | 8 | 8 |
| Straight (attracted to people of the opposite gender) | 97 | 41 | 35 | 40 | 47 |
| Prefer to self-describe<sup>3</sup> | 10 | 4 | 6 | 3 | 4 |
| Prefer not to say | 9 | 4 | 5 | 5 | 1 |

1: Agender, Genderfluid, Genderqueer, Non-binary, Non-binary AFAB, She/They, Trans female to male, Transmasculine
2: Portuguese
3: Not straight, Not sure, Omnisexual, Pansexual, Queer and/or lesbian, Sapphic, Unlabelled

#### Self-harm thoughts and behaviours

Most participants had engaged in non-suicidal self-harm (99%), had thoughts that life was not worth living (i.e., passive suicidal thoughts) (97%), and had experienced suicidal ideations (i.e., active suicidal thoughts) (93%) during their lifetime. Just over half the sample (56%) had made a suicide attempt. Figures 1 and 2 display information about the recency and past year frequency of self-harm thoughts and behaviours in the sample.

**Figure 1.**
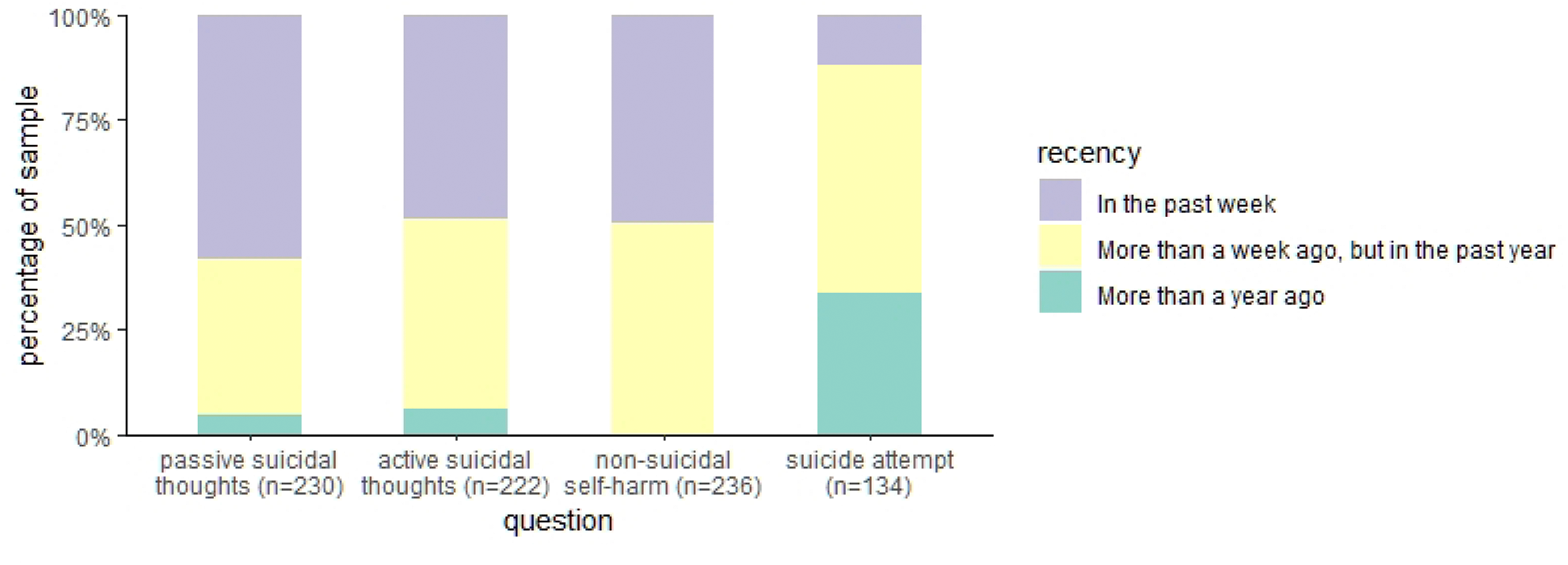
Recency of suicidal thoughts and self-harm behaviour

**Figure 2.**
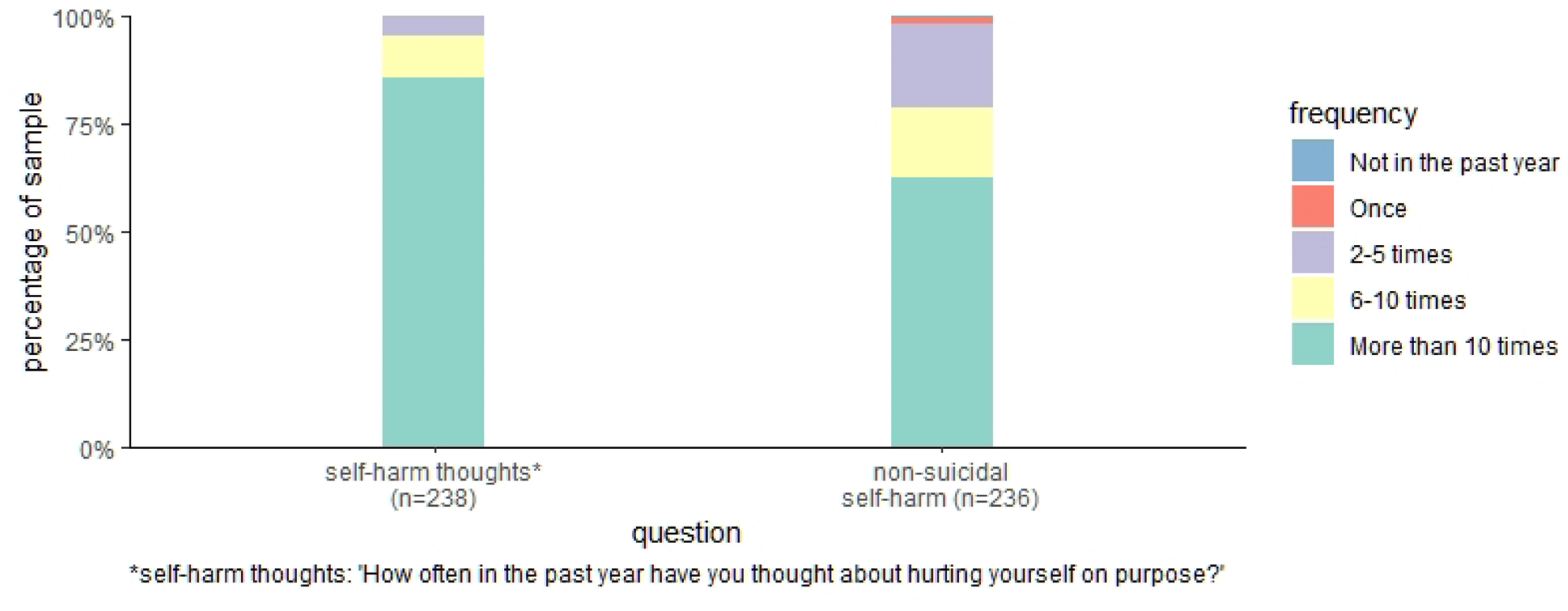
Frequency of self-harm thoughts and behaviour (in the past year)

In terms of type of self-harm methods used, 43% reported swallowing pills or something poisonous, 45% reported burning themselves, and 95% reported cutting themselves. Using another method not listed was reported by 45% of participants. These were screened by JF and BM and then categorised (for full details, please refer to Table S3 in Appendix S2). Behaviours not considered to be self-harm (for example tattooing, eating behaviours or indirect self-harm such as unsafe sex) were removed, however all participants reporting these behaviours also reported at least one other self-harm method, and so no participants were removed from the analysis. Most participants (41%) reported having used two self-harm methods during their lifetime, with 21%, 25% and 13% reporting one, three and four or more methods respectively.

#### Help-seeking history

When reflecting on help-seeking over their lifetime, seeking help from no one was reported by 16% of participants (n=39). For a breakdown of the prevalence of accessing each source of informal and formal help, please refer to Table S4 in Appendix S2.

### Outcome measures

Descriptive statistics are reported for the five outcome measures by experimental condition in Table 2.

**Table 2.**
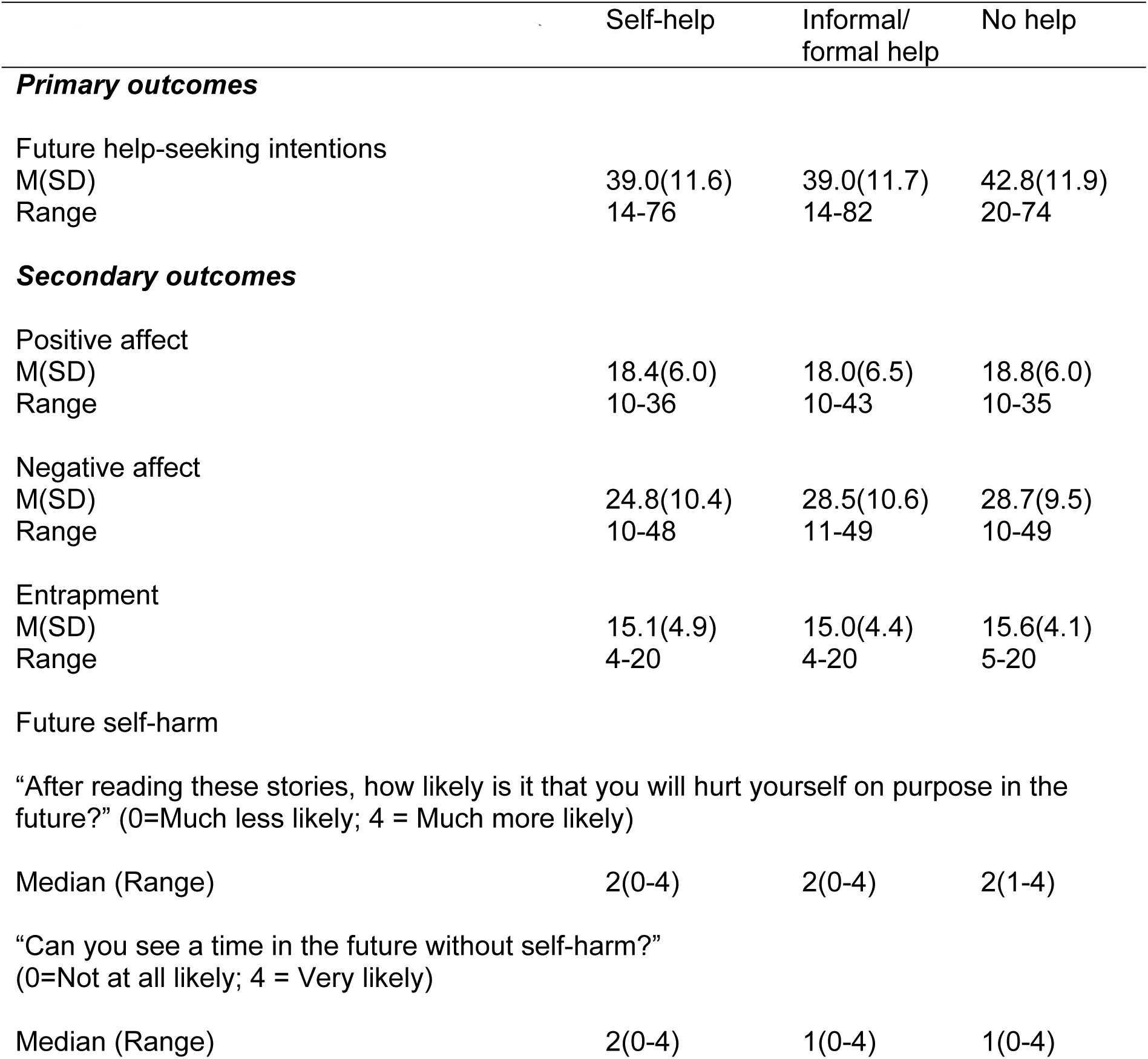
Descriptive statistics for outcomes.

| Table 2. Descriptive statistics for outcomes | Self-help | Informal/<br>formal help | No help |
| --- | --- | --- | --- |
| **Primary outcomes** |  |  |  |
| Future help-seeking intentions |  |  |  |
| M(SD) | 39.0(11.6) | 39.0(11.7) | 42.8(11.9) |
| Range | 14-76 | 14-82 | 20-74 |
| **Secondary outcomes** |  |  |  |
| Positive affect |  |  |  |
| M(SD) | 18.4(6.0) | 18.0(6.5) | 18.8(6.0) |
| Range | 10-36 | 10-43 | 10-35 |
| Negative affect |  |  |  |
| M(SD) | 24.8(10.4) | 28.5(10.6) | 28.7(9.5) |
| Range | 10-48 | 11-49 | 10-49 |
| Entrapment |  |  |  |
| M(SD) | 15.1(4.9) | 15.0(4.4) | 15.6(4.1) |
| Range | 4-20 | 4-20 | 5-20 |
| Future self-harm |  |  |  |
| “After reading these stories, how likely is it that you will hurt yourself on purpose in the future?” (0=Much less likely; 4 = Much more likely) |  |  |  |
| Median (Range) | 2(0-4) | 2(0-4) | 2(1-4) |
| “Can you see a time in the future without self-harm?” (0=Not at all likely; 4 = Very likely) |  |  |  |
| Median (Range) | 2(0-4) | 1(0-4) | 1(0-4) |

#### Future help-seeking

Due to the skewed nature of the data, future help-seeking data was transformed using a square root transformation. There was weak to moderate evidence for an effect of story type on future help-seeking intentions F(2,235) = 2.8, p = .06, η^2^ = .02. Adjusting for age, gender, and help-seeking history provided stronger evidence for an effect of story type on future-help seeking, F(2,230) = 3.5, p = .03, η^2^ = .21 (the full results of the model using untransformed and untransformed data can be found in Tables S5 and S6 of Appendix S3). However, none of the pairwise comparisons (Tukey’s HSD on the unadjusted means) suggested that there were differences in future help-seeking among the three groups. The impact of story type on future help-seeking is illustrated by Figure 3 (using untransformed data).

**Figure 3.**
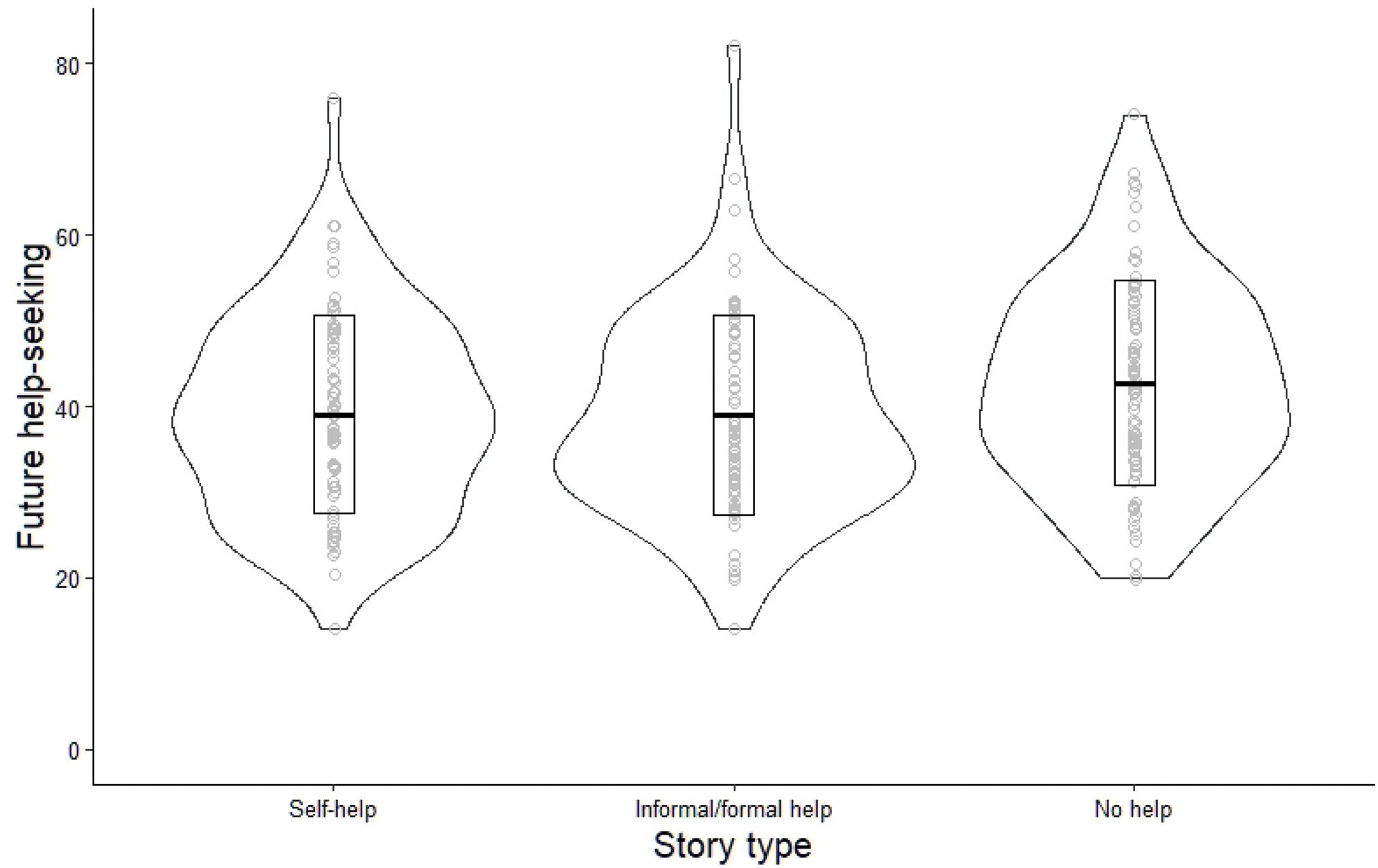
Impact of story type on future help-seeking

An ANCOVA was conducted to determine whether self-harm history (recency of self-harm) interacted with story type to influence future help-seeking. Evidence for this interaction was not found, F(2,232) = 0.57, p = 0.57. An ANCOVA was conducted to explore if age interacted with story type to influence future help-seeking. Evidence for this interaction was not found, F(2,232) = 1.88, p =.15. However, the former only achieved 15% power, and the latter 41% power.

#### Affect, entrapment, future self-harm

Due to the skewed nature of the data, attempts to transform all secondary outcomes were made but were unsuccessful. Therefore, the following analyses were conducted using untransformed data.

We found no evidence for an effect of story type on positive affect, F(2,235) = .32, p = .73, η^2^ = .002 or entrapment, F(2,235) = .33, p = .72, η^2^ = .003. There was also no evidence for an effect of story type on future self-harm (Likelihood of self-harm after reading stories, F(2,235) = 2.40, p = .09, η^2^ = .02; Likelihood of a future without self-harm, F(2,235) = 0.94, p = .39, η^2^ = .008). The pattern of results was unchanged following adjustment for age, gender, and help-seeking history (the full results of the model can be found in Tables S7, S9, S10, and S11 of Appendix S3).

There was evidence for an effect of story type on negative affect, F(2,235) =3.83, p = .02, η^2^ = .03. Adjusting for age, gender, and help-seeking history, increased the strength of association between story type and negative affect F(2,230) = 4.02, p = .02, η^2^ = .10 (the full results of the model can be found in Table S8 of Appendix S3). Pairwise comparisons (Tukey’s HSD on the unadjusted means) demonstrated that participants in the “Self-help” condition reported lower levels of negative affect after reading the stories compared to participants in the “No help” condition (Mean difference = -3.97, 95% CI -7.72 to -0.22, p = .04) and the “Informal/formal” help condition (Mean difference = -3.70, 95% CI -7.55 to 0.14, p =.06). Figure 4 presents the impact of story type on negative impact.

**Figure 4.**
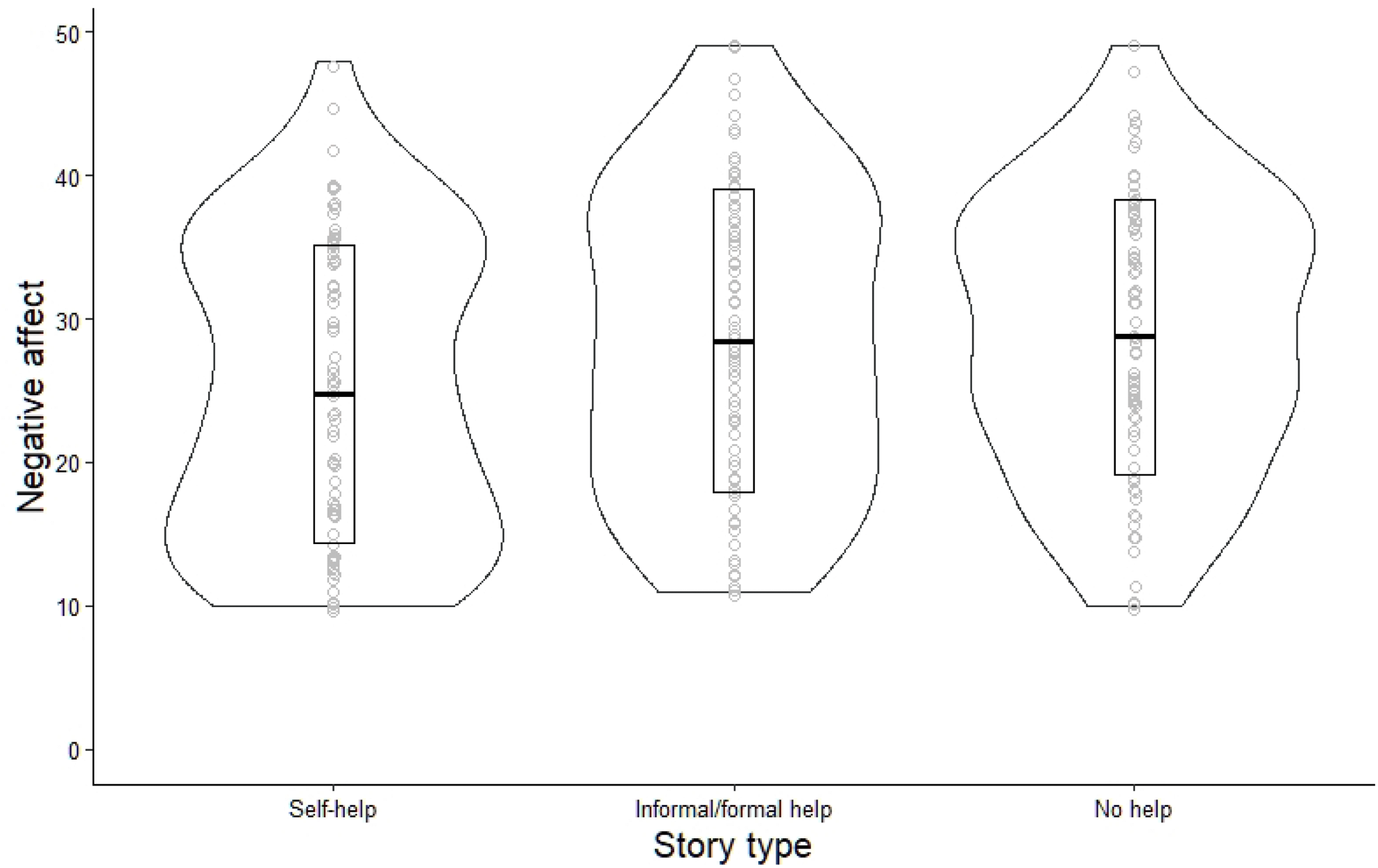
Impact of story type on negative affect

An ANCOVA was conducted to explore if self-harm history interacted with story type to influence negative affect. Although there was strong evidence for an effect of self-harm history on negative affect, F(1,232) = 28.94, p <.001, evidence for an interaction with story type was not found, F(2,232) = 0.94, p = 0.39. An ANCOVA was also conducted to explore if age interacted with story type to influence negative affect. Although there was strong evidence for an effect on age on negative affect, F(1,232) = 16.85, p < .001, evidence for an interaction was not found, F(2,232) = 1.06, p =.35. However, the former only achieved 22% power, and the latter 26% power.

### Open-ended feedback on stories

Eighty-eight percent of the sample provided written feedback on the stories. Limitations and advantages of the lived experience stories, categorised by the type of story, are reported in Tables S12 and S13 in Appendix S4. Below, we have reported noticeable differences in the themes across conditions.

#### Differences in perceived limitations of the stories across conditions

##### The recovery messaging was problematic

Within the comments, messages of hope, positivity, and recovery were described as annoying, frustrating, and overwhelming. Some participants expressed dislike towards references to counting the number of days without self-harm. Other comments referred to the difficulty in looking to the future as that felt dismissive and more challenging than taking it day-by-day. A participant explained,

> “I don’t care if I’m fine by the time I’m 21. I’ve ruined my teenage years with this, and I can’t get that back. Yeah, I hope for a happier future, but when people say that everything is fine because they recovered (…) they’re ignoring how horrific the emotions were when they were struggling. Recovery doesn’t magically erase the past”. – Participant in Informal/Formal Help condition

Another participant expressed frustration that the narrators in the stories seemed to be working towards recovery due to feeling stigmatised and pressured to conform, rather than of their own accord. The “No help” condition stands out in that more participants appeared to struggle with the inclusion of recovery messaging compared to the other story conditions. This may be because the recovery message appeared without a clear indication of how recovery occurred as comments reflected the following sentiments,

> “Sometimes words can get lost e.g., “things get better”. The term has been used so many times that I have almost become desensitized to it and what it means.” - Participant in No Help condition “Doesn’t say how to get better.” - Participant in No Help condition

##### The stories were unrelatable

While some participants in all conditions commented on the stories being unrelatable, this was less pronounced in the “No help” condition. This is plausibly a result of the stories in this condition providing less concrete detail about the narrator’s experience and thus there being less to be able to relate to. Stories in the “Self-help” condition were criticised more than stories in the other conditions for neglecting to acknowledge that “one size does not fit all”. This was mostly reflected in the criticism that the stories were too specific, usually because of a focus on younger ages. However, some participants in the “Self-help” condition agreed with the sentiment that “the people in the stories seemed to have a better idea of why they started self-harming and why they continued to do it, which I don’t have”.

The most prevalent reasons for why the stories were unrelatable referred to the stories having unrealistic portrayals of self-harm, and instilling feelings of competition and thus feelings of isolation in readers. They also commented that they felt pressure to have “stronger urges” and “scars” like the narrators’ had. Participants in all conditions found the stories unrealistic to a similar degree, however more references about the stories seeming sanitised or inauthentic were made by participants in the “Self-help” condition. Participants in the “Self-help” and “Informal/formal help” commented on the stories making them feel isolated much more than those in the “No help” condition. Several participants emphasised that there is a very competitive nature to self-harm at both extremes:

> “It can sometimes make you feel as if you are not bad enough to deserve help”. – Participant in “Self-help” condition

> “All people who seemed to be able to stop with help. It made me feel bad for still struggling.” – Participant in “Informal/formal help” condition

##### The specific support mentioned in the stories (or lack thereof) was problematic

The stories in the “Self-help” condition were criticised for not mentioning professional or medical support. All conditions were criticised for offering non-practical advice. This was more predominant in the “Self-help” condition, followed by “Informal/formal help” condition, and much less in the “No help” condition. The type of self-help methods mentioned were mainly criticised for being methods that readers had already tried and found unsuccessful in aiding recovery; there were also a couple of comments criticising the focus on short-term coping mechanisms, instead of long-term recovery.

> “After trying the methods they said helped them, I eventually slipped back to self harming. I don’t know what else to do.” -Participant in “Self-help” condition

> “They didn’t give much detail on what actually helped them other than immediate coping mechanisms.” - Participant in “Self-help” condition

The type of informal/formal help methods mentioned were mainly criticised as participants felt the stories made accessing those kinds of support sound much easier than it is. For example, a participant stated,

> “Sometimes it may not be as easy to access support services, whilst barriers to access are likely to vary, it felt like the people in the stories were able to somewhat easily access support - the way it should be I guess.” - Participant in “Informal/formal help”condition

Another stated,

> “I’m a first-generation immigrant, mental health was never something to talk about in my household because of cultural reasons. If I had gone to my parents, I know I wouldn’t get the same sympathy just frustration.” - Participant in “Informal/formal help” condition

This made readers who did not have supportive friends/family or have struggled to access formal support, feel isolated and distressed,

> “I guess it makes me sad to hear about people having their boyfriend, friends and family around to help them when I feel like I have no one.” - Participant in “Informal/formal help” condition

#### Differences in perceived advantages of the stories across conditions

While the “Informal/formal help” condition did not stand out in the numbers of criticisms received, participants in this condition did seem to have fewer positive things to say about the stories. This was apparent in relation to comments about the stories having a realistic but hopeful outlook of recovery and comments about the authenticity of the stories.

##### The stories included a realistic but hopeful outlook of recovery

Participants in all conditions appreciated hearing the stories from those who recovered, and particularly the emphasis on recovery being a nonlinear process that takes time. Only the “Self-help” stories were praised for highlighting that the path to recovery is not a one-size-fits-all approach, and that readers will find different coping mechanisms and recovery strategies helpful. As mentioned earlier, participants in the “Self-help” condition were also more likely to criticize the stories for failing to do exactly this. Only the stories in the “Self-help” and “No help” conditions were praised for the discussion of relapses, despite these themes appearing in all conditions.

##### The stories were relatable

The number of positive comments about the relatability of the stories were similar across conditions. However, there were some differences among the conditions when looking at the subthemes of relatability. Although positive comments about the authenticity of the stories were made about all of the stories, this was more prevalent in the “Self-help” and “No help” conditions. All of the stories were commended for accurately portraying the emotions experienced in self-harm, but the stories in the “Self-help” and “No help” conditions were further commended for relaying “lived experience” and seeming “honest and truthful”. One participant explained, that,

> “It can be hard to accept advice from people that don’t fully understand or appreciate just how dark the thoughts can be and how strong these urges and negative thoughts can take over your mind”. – Participant in “No help” condition

We note that there are inconsistences between the unrelatable and relatable themes which we will elaborate on in the discussion.

##### The stories were supportive

Unsurprisingly, participants in the “No help” condition had fewer positive remarks about the theme of support compared to participants in the conditions that focused on help-seeking. However, this difference was not as substantial as some participants in this condition felt that hearing others’ stories reminded them that they were not alone. It is also worth noting that this feeling of ‘camaraderie’ was reported a similar number of times across conditions. Other aspects that fell under the theme of support seen in the other conditions were: Participants in the “Self-help” condition appreciated hearing what specific methods worked for others as it gave them hope generally but also ideas about new coping mechanisms to try. Participants in the “Informal/formal help” condition appreciated the emphasis on the importance of accessing support for recovery. The stories highlighted to them how helpful it was to have both informal and formal help at the start but also throughout recovery. The stories could help to instill hope and confidence,

> “It does get better once you find the right support. There are people who do care about you, you just don’t know it.” - Participant in “Informal/formal help” condition

While some participants who had negative or no help-seeking experiences in the past found it difficult to read about others having success, some participants found it reassuring to hear about other people successfully accessing support as there is concern that accessibility can be restricted.

## Discussion

### Main findings

In this study of individuals with recent self-harm experience, we were unable to find clear evidence of an association between the type of content of lived experience stories and attitudes towards future help-seeking. One potential explanation for why we did not see an increase in help-seeking intentions after reading a story that emphasizes help-seeking might be due to the relatability of the stories. The open-ended feedback suggested that the stories that mentioned help-seeking were seen as less relatable than the stories that did not mention help-seeking. This demonstrates that it is not only the characteristics of the narrator (e.g., age, gender) that the reader might not relate to. They might also struggle to relate to the actions and consequences of help-seeking experienced by the narrator. Firstly, about 13% of participants reported no history of help-seeking. Others who recounted negative experiences when they sought help (e.g., ineffective self-help strategies, negative reactions from family and friends, inaccessible formal support) reported feeling distressed (i.e., feeling like a failure, alone, and hopeless) when they read about others’ positive experiences. Further, only the stories that mentioned help-seeking seemed to evoke a feeling that the reader’s situation was less serious than the narrator’s. As perceived need is an important component of help-seeking decision making, (Mojtabai et al., 2002), it is feasible that the stories that mentioned help-seeking set a benchmark for when help-seeking is needed. In our focus group study, we similarly found that being able to identify with the narrator of a lived experience story – made more challenging by including specific details of age or circumstances – was “an important factor in encouraging help-seeking through the recognition of need in others and thus oneself” (23).

Findings need to be interpreted in light of some limitations. Firstly, we did not measure help-seeking intentions before and after the exposure so we cannot determine if there was an overall increase across all participants. Secondly, the study was powered to detect a main effect of type of lived experience story, and tests of interactions were exploratory and underpowered. These post-hoc findings therefore need to be interpreted with caution, and larger-scale research is needed to understand the role of moderators, such as self-harm history and age. Thirdly, the sample was self-selecting and its possible they may not be representative of the wider population.

It is worth noting that some of the themes that emerged from the open-ended feedback questions contradicted one another. For example, the self-help stories seemed to arouse thoughts about self-harm and recovery being very individualistic experiences; but they were both praised for doing this well and critiqued for doing it poorly. Similarly, the same aspect of a story could be a positive that made the story more relatable for one participant, and a negative addition that made the story less relatable for another. While some attempts were made to reconcile contradictions like this when interpreting the data, we were conservative in our approach. Firstly, because we felt that the contradictions were the result of a broad focus. Feedback was collected from a large number of individuals and that feedback could be directed towards various aspects of the stories. Secondly, the contradictions in themselves may be meaningful, as they indicate how individualistic the experience of self-harm and recovery can be, and stress that a delicate balance needs to be found in these stories, if hoping to apply them broadly.

Overall, there was broad consistency between the themes that emerged from the open-ended feedback collected in this study and those that emerged during the focus group discussions with 5% of the original sample (23). Both studies highlighted the importance for lived experience stories to be relatable, but that a careful balance had to be struck between specificity and ambiguity. Both studies also highlighted the importance for lived experience stories to feel authentic (avoiding both dramatisation and sanitisation) and that authenticity is a key factor in how relatable a story is to the reader. Previous research on mental health recovery narratives also stress the importance of authenticity to readers (36,37). While the template stories were written by those with lived experience, it was up to the research team’s discretion about how to modify them for the purpose of this study, and this might have impacted the relatability, or other aspects, of the stories. Therefore, we suggest utilising a co-design approach in future work.

In line with the findings of Winstone et al (2023), this study also highlighted the importance for lived experience stories to offer practical advice and to portray the journey to recovery realistically. This finding might be specific to self-harm recovery, as another study which more broadly focused on participants with any mental health concern and excluded narratives that described harmful behaviour (e.g., self-harm) found participants reported a preference for narratives about upward trajectories towards recovery over narratives that focused on only snapshots of the journey or non-linear journeys (36).

Results from the focus groups suggest that reading the stories had a generally positive impact on mood (Winstone et al., 2023). We did not measure baseline mood in the present study, and we cannot assume that if we did that the results would support the qualitative findings, as the focus group sample might have been prone to sampling bias since it is possible that participants who felt the stories negatively impacted their mood were less likely to enrol in the follow-up qualitative study. There was however strong evidence that reading stories about self-help strategies led to lower prevalence of negative emotions, compared to reading stories about seeking help from others or which made no mention of help-seeking. The open-ended feedback suggested that reading about self-help strategies that were successful for others left some readers feeling hopeful that recovery might be possible for them too. It also gave them ideas of new strategies to try, inspiring them to act towards achieving their own recovery. It is possible that these positives may have increased their self-efficacy (38) to recover, thereby increasing their mood and decreasing feelings of needing to rely on others for help. While this interpretation is speculative, further investigation is warranted since an unintended negative consequence may be a decreased likelihood of seeking help from others.

The results from this study suggest that readers might react more negatively to themes of recovery in the absence of practical advice on how to recover. The results from this study also add the caveat that it would be beneficial to mention various help-seeking strategies within the same story. This might prevent the reader from feeling demoralised (and avoid seeking help) after reading about someone else successfully obtaining help via a strategy that was unsuccessful for the reader or is a strategy that they are unwilling to try.

Our findings, consistent with the findings of Winstone et al (2023), demonstrate that lived experience stories can help readers to “feel validated and help them to better understand their own feelings and experiences” and manage “feelings of isolation, knowing that others are experiencing similar feelings, and engaging in self-harm as a coping mechanism” (Winstone et al., 2023, p. 7). However, the results from both studies also highlight that reading about others’ experiences can sometimes foster a sense of competition, and thereby increase feelings of isolation. Previous research has also highlighted the competitive element of self-harm (19,39,40) and found that individuals sometimes purposefully seek out material which instills feelings of competition when experiencing self-harm urges (39).

One way to prevent lived experience stories from triggering a sense of competition in readers is by avoiding words such as “clean” or other language that might cause readers to feel stigmatized or under pressure to recover. In addition, it might be helpful to explicitly refer, in lived experience stories, to the competitive nature of self-harm. For example, narrators can emphasize that they are relaying only one out of many stories (that not everyone’s self-harm will have the same causes, that not everyone will experience it in the same way, and that not everyone will recover in the same way) but that everyone’s experience is valid. Winstone et al (2023) further concluded that avoiding numeric references within a story, such as number of days without self-harm, might help to reduce competition.

The lived experience stories under study were carefully selected to be recovery-oriented and exclude details that might be triggering for someone who self-harms (e.g., detailed descriptions of self-harm methods). Despite these attempts, the results demonstrated that some participants (across all conditions) reported emotional distress as a result of reading the stories. Some participants were negatively impacted by mention of the specific self-harm method used (i.e., cutting). While lived experience stories on forums often include descriptive detail about methods and injuries (41), those hosted on more formal websites (e.g., run by charities and healthcare organisations) usually exclude these kinds of details (22). However, some participants were negatively impacted by other details that might be found in stories hosted on formal websites: hearing about others struggling with self-harm, being reminded of how they harm themselves, or how one’s self-harm might impact their loved ones. It was also reported that reading about others still struggling with self-harm made readers feel hopeless that their situation would ever meaningfully improve. Therefore, future research, utilizing a co-design approach, is needed to identify elements that make a story harmful.

### Conclusions

The findings taken together illustrate that the impact of lived experience stories on individuals with a recent history of self-harm is complicated. While the inclusion of help-seeking in a story can help readers to feel validated, less isolated, less helpless, and more hopeful that recovery is possible for them, there is a fine line to tread. The results indicate that the experience of self-harm and the pathway to recovery is very personal, so too many personal details about either experience can make the stories feel unrelatable. However, there are certain shared experiences of self-harm that can be included to ensure a lived experience story feels authentic. Therefore, the results suggest that stories should aim to foster both a sense of individuality as well as community. For example, stories should focus less on the characteristics of narrators, but more on their emotions and experience of self-harm to which readers can be more easily relate. Stories should also propose multiple strategies for achieving recovery, and emphasise that recovery will take time, and that there may be setbacks along the way. Future experimental work is needed to confirm if these adaptations to lived experience stories about help-seeking can motivate individuals to seek-help for their self-harm.

## Data Availability

Open access statement and link to data.bris repository to be added on manuscript upon acceptance.

## Acknowledgments

We would like to thank Jenna Selvey for assisting with the coding of the open-ended feedback on the lived-experience stories.

## References

1. Boyce N. Pilots of the future: suicide prevention and the internet. The Lancet [Internet]. 2010 Dec 4 [cited 2022 Apr 8];376(9756):1889–90. Available from: http://www.thelancet.com/article/S014067361062199X/fulltext

2. Brennan C, Saraiva S, Mitchell E, Melia R, Campbell L, King N, et al. Self-harm and suicidal content online, harmful or helpful? A systematic review of the recent evidence. J Public Ment Health [Internet]. 2022 [cited 2024 Apr 17];21(1). Available from: https://www.emerald.com/insight/content/doi/10.1108/JPMH-09-2021-0118/full/html

3. Baker D, Fortune S. Understanding self-harm and suicide websites: a qualitative interview study of young adult website users. Crisis [Internet]. 2008 [cited 2022 Apr 13];29(3):118–22. Available from: https://pubmed.ncbi.nlm.nih.gov/18714907/

4. Harris IM, Roberts LM. Exploring the use and effects of deliberate self-harm websites: an Internet-based study. J Med Internet Res [Internet]. 2013 [cited 2022 Apr 13];15(12). Available from: https://pubmed.ncbi.nlm.nih.gov/24362563/

5. Shaw LH, Gant LM. In defense of the internet: the relationship between Internet communication and depression, loneliness, self-esteem, and perceived social support. Cyberpsychol Behav [Internet]. 2002 [cited 2022 Apr 13];5(2):157–71. Available from: https://pubmed.ncbi.nlm.nih.gov/12025883/

6. Whitlock JL, Powers JL, Eckenrode J. The virtual cutting edge: the internet and adolescent self-injury. Dev Psychol [Internet]. 2006 May [cited 2022 Apr 13];42(3):407–17. Available from: https://pubmed.ncbi.nlm.nih.gov/16756433/

7. Lai MH, Maniam T, Chan LF, Ravindran A v. Caught in the web: a review of web-based suicide prevention. J Med Internet Res [Internet]. 2014 [cited 2022 Apr 13];16(1). Available from: https://pubmed.ncbi.nlm.nih.gov/24472876/

8. Luxton DD, June JD, Kinn JT. Technology-Based Suicide Prevention: Current Applications and Future Directions. https://home.liebertpub.com/tmj [Internet]. 2011 Feb 6 [cited 2022 Apr 13];17(1):50–4. Available from: https://www.liebertpub.com/doi/abs/10.1089/tmj.2010.0091

9. Robinson J, Cox G, Bailey E, Hetrick S, Rodrigues M, Fisher S, et al. Social media and suicide prevention: a systematic review. Early Interv Psychiatry [Internet]. 2016 Apr 1 [cited 2022 Apr 13];10(2):103–21. Available from: https://pubmed.ncbi.nlm.nih.gov/25702826/

10. Scherr S, Reinemann C. First do no harm: Cross-sectional and longitudinal evidence for the impact of individual suicidality on the use of online health forums and support groups. Comput Human Behav [Internet]. 2016 Aug 1 [cited 2022 Apr 13];61:80–8. Available from: https://www.sciencedirect.com/science/article/pii/S0747563216301625

11. Frost M, Casey L. Who Seeks Help Online for Self-Injury? 10.1080/1381111820151004470 [Internet]. 2016 Jan 2 [cited 2022 Jul 20];20(1):69–79. Available from: https://www.tandfonline.com/doi/abs/10.1080/13811118.2015.1004470

12. Mars B, Heron J, Biddle L, Donovan JL, Holley R, Piper M, et al. Exposure to, and searching for, information about suicide and self-harm on the Internet: Prevalence and predictors in a population based cohort of young adults. J Affect Disord [Internet]. 2015 Oct 1 [cited 2022 Apr 8];185:239. Available from: /pmc/articles/PMC4550475/

13. Biddle L, Derges J, Goldsmith C, Donovan JL, Gunnell D. Online help for people with suicidal thoughts provided by charities and healthcare organisations: a qualitative study of users’ perceptions. Soc Psychiatry Psychiatr Epidemiol [Internet]. 2020 Sep 1 [cited 2022 Jul 20];55(9):1157–66. Available from: https://link.springer.com/article/10.1007/s00127-020-01852-6

14. Greidanus E, Everall RD. Helper therapy in an online suicide prevention community. Br J Guid Counc [Internet]. 2010 [cited 2024 Apr 17];38(2). Available from: https://www.tandfonline.com/doi/full/10.1080/03069881003600991

15. Padmanathan P, Biddle L, Carroll R, Derges J, Potokar J, Gunnell D. Suicide and Self-Harm Related Internet Use: A Cross-Sectional Study and Clinician Focus Groups. Crisis [Internet]. 2018 [cited 2022 Apr 8];39(6):469. Available from: /pmc/articles/PMC6263311/

16. Biddle L, Derges J, Goldsmith C, Donovan JL, Gunnell D. Using the internet for suicide-related purposes: Contrasting findings from young people in the community and self-harm patients admitted to hospital. PLoS One [Internet]. 2018 May 1 [cited 2022 Apr 8];13(5):e0197712. Available from: https://journals.plos.org/plosone/article?id=10.1371/journal.pone.0197712

17. Biddle L, Donovan J, Hawton K, Kapur N, Gunnell D. Public Health: Suicide and the internet. BMJ: British Medical Journal [Internet]. 2008 Apr 4 [cited 2022 Jul 20];336(7648):800. Available from: /pmc/articles/PMC2292278/

18. Lewis SP, Seko Y. A Double-Edged Sword: A Review of Benefits and Risks of Online Nonsuicidal Self-Injury Activities. J Clin Psychol [Internet]. 2016 [cited 2024 Apr 17];72(3). Available from: https://pubmed.ncbi.nlm.nih.gov/26613372/

19. Susi K, Glover-Ford F, Stewart A, Knowles Bevis R, Hawton K. Research Review: Viewing self-harm images on the internet and social media platforms: systematic review of the impact and associated psychological mechanisms. Journal of Child Psychology and Psychiatry [Internet]. 2023 Aug 1 [cited 2023 Jul 18];64(8):1115–39. Available from: https://onlinelibrary.wiley.com/doi/full/10.1111/jcpp.13754

20. McDermott E, Roen K, Piela A. Hard-to-reach youth online: Methodological advances in self-harm research. Sexuality Research and Social Policy [Internet]. 2013 Jun 24 [cited 2022 Jul 20];10(2):125–34. Available from: https://link.springer.com/article/10.1007/s13178-012-0108-z

21. Daine K, Hawton K, Singaravelu V, Stewart A, Simkin S, Montgomery P. The Power of the Web: A Systematic Review of Studies of the Influence of the Internet on Self-Harm and Suicide in Young People. PLoS One [Internet]. 2013 [cited 2022 Jul 20];8(10):e77555. Available from: https://journals.plos.org/plosone/article?id=10.1371/journal.pone.0077555

22. Cohen R, Rifkin-Zybutz R, Moran P, Biddle L. Web-based support services to help prevent suicide in young people and students: A mixed-methods, user-informed review of characteristics and effective elements. Health Soc Care Community [Internet]. 2022 May 6 [cited 2023 Sep 27]; Available from: https://www.ncbi.nlm.nih.gov/pmc/articles/PMC10084127/

23. Winstone L, Mars B, Ferrar J, Moran P, Penton-Voak I, Grace L, et al. Investigating How People Who Self-harm Evaluate Web-Based Lived Experience Stories: Focus Group Study. JMIR Ment Health [Internet]. 2023 [cited 2023 Jun 14];10. Available from: https://pubmed.ncbi.nlm.nih.gov/36719729/

24. Bechmann A, Lomborg S. Mapping actor roles in social media: Different perspectives on value creation in theories of user participation. New Media Soc [Internet]. 2013 [cited 2024 Apr 17];15(5). Available from: https://journals.sagepub.com/doi/abs/10.1177/1461444812462853?journalCode=nms a

25. Bailey ER, Matz SC, Youyou W, Iyengar SS. Authentic self-expression on social media is associated with greater subjective well-being. Nature Communications 2020 11:1 [Internet]. 2020 Oct 6 [cited 2023 Aug 16];11(1):1–9. Available from: https://www.nature.com/articles/s41467-020-18539-w

26. O’Connor RC, Portzky G. The relationship between entrapment and suicidal behavior through the lens of the integrated motivational–volitional model of suicidal behavior. Curr Opin Psychol [Internet]. 2018 Aug 1 [cited 2023 Jul 18];22:12–7. Available from: https://pubmed.ncbi.nlm.nih.gov/30122271/

27. Willliams JMG. Suicide and Attempted Suicide: Understanding the Cry of Pain [Internet]. 2nd ed. London: Penguin Books; 2002 [cited 2023 Jul 18]. Available from: https://books.google.co.uk/books?hl=en&lr=&id=s-FdG3U0rwEC&oi=fnd&pg=PR6&ots=mQaLs0-HkT&sig=TdXZs5do0GB9V363MpKTiS2glQM&redir_esc=y#v=onepage&q=entrapment&f=false

28. Faul F, Erdfelder E, Buchner A, Lang AG. Statistical power analyses using G*Power 3.1: Tests for correlation and regression analyses. Behav Res Methods [Internet]. 2009 [cited 2022 Mar 9];41(4):1149–60. Available from: https://www.psychologie.hhu.de/fileadmin/redaktion/Fakultaeten/Mathematisch-Naturwissenschaftliche_Fakultaet/Psychologie/AAP/gpower/GPower31-BRM-Paper.pdf

29. Anwyl-Irvine AL, Massonnié J, Flitton A, Kirkham N, Evershed JK. Gorilla in our midst: An online behavioral experiment builder. Behav Res Methods [Internet]. 2020 Feb 1 [cited 2022 Feb 4];52(1):388–407. Available from: https://link.springer.com/article/10.3758/s13428-019-01237-x

30. Rickwood D, Deane FP, Wilson CJ, Ciarrochi J. Young people’s help-seeking for mental health problems. Australian e-Journal for the Advancement of Mental Health [Internet]. 2005 [cited 2022 Feb 15];4(3). Available from: www.auseinet.com/journal/vol4iss3suppl/rickwood.pdfwww.auseinet.com/journal

31. Rickwood DJ, Braithwaite VA. Social-psychological factors affecting help-seeking for emotional problems. Soc Sci Med [Internet]. 1994 Aug 1 [cited 2022 Jan 31];39(4):563–72. Available from: https://pubmed.ncbi.nlm.nih.gov/7973856/

32. Wilson MMG, Morley JE. Impaired cognitive function and mental performance in mild dehydration. Eur J Clin Nutr [Internet]. 2003 Dec [cited 2015 May 5];57 Suppl 2(S2):S24–9. Available from: 10.1038/sj.ejcn.1601898

33. Watson D, Clark LA, Tellegen A. Development and validation of brief measures of positive and negative affect: the PANAS scales. J Pers Soc Psychol [Internet]. 1988 [cited 2022 Feb 1];54(6):1063–70. Available from: https://pubmed.ncbi.nlm.nih.gov/3397865/

34. de Beurs D, Cleare S, Wetherall K, Eschle-Byrne S, Ferguson E, B O’Connor D, et al. Entrapment and suicide risk: The development of the 4-item Entrapment Scale Short-Form (E-SF). Psychiatry Res [Internet]. 2020 Feb 1 [cited 2022 Feb 2];284. Available from: https://pubmed.ncbi.nlm.nih.gov/31945600/

35. Oppenheimer DM, Meyvis T, Davidenko N. Instructional manipulation checks: Detecting satisficing to increase statistical power. J Exp Soc Psychol [Internet]. 2009 [cited 2017 Feb 22];45:867–72. Available from: https://www.sciencedirect.com/science/article/pii/S0022103109000766

36. Ng F, Charles A, Pollock K, Rennick-Egglestone S, Cuijpers P, Gillard S, et al. The mechanisms and processes of connection: Developing a causal chain model capturing impacts of receiving recorded mental health recovery narratives. BMC Psychiatry [Internet]. 2019 Dec 21 [cited 2023 Jul 18];19(1):1–15. Available from: https://bmcpsychiatry.biomedcentral.com/articles/10.1186/s12888-019-2405-z

37. Rennick-Egglestone S, Ramsay A, McGranahan R, Llewellyn-Beardsley J, Hui A, Pollock K, et al. The impact of mental health recovery narratives on recipients experiencing mental health problems: Qualitative analysis and change model. PLoS One [Internet]. 2019 Dec 1 [cited 2023 Jul 18];14(12). Available from: https://pubmed.ncbi.nlm.nih.gov/31834902/

38. Bandura A. Self-efficacy: Toward a unifying theory of behavioral change. Psychol Rev [Internet]. 1977 [cited 2024 Apr 17];84(2). Available from: https://www.sciencedirect.com/science/article/pii/0146640278900024

39. Murray CD, Fox J. Do Internet self-harm discussion groups alleviate or exacerbate self-harming behaviour? 10.5172/jamh53225 [Internet]. 2014 Jan [cited 2023 Jul 18];5(3):225–33. Available from: https://www.tandfonline.com/doi/abs/10.5172/jamh.5.3.225

40. Seif NA, Bastien RJB, Wang B, Davies J, Isaken M, Ball E, et al. Effectiveness, acceptability and potential harms of peer support for self-harm in non-clinical settings: systematic review. BJPsych Open [Internet]. 2022 Jan [cited 2023 Jul 18];8(1). Available from: /pmc/articles/PMC8811789/

41. Recovery Your Life [Internet]. [cited 2023 Sep 27]. Self-Harm Discussion and Support. Available from: https://www.recoveryourlife.com/forum/forumdisplay.php?f=29

